# Individuals who were mildly symptomatic following infection with SARS-CoV-2 B.1.1.28 have neutralizing antibodies to the P.1 variant

**DOI:** 10.1101/2021.05.11.21256908

**Authors:** Maria Cassia Mendes-Correa, Lucy S. Villas-Boas, Ana Luiza Bierrenbach, Anderson de Paula, Tania Regina Tozetto-Mendoza, Fabio E. Leal, Wilton Freire, Heuder Gustavo Oliveira Paiao, Andrea B C Ferraz, Steven S. Witkin

**Affiliations:** Faculdade de Medicina da Universidade de Sao Paulo-Laboratorio de Investigacao Medica em Virologia (LIM52)-Instituto de Medicina Tropical de Sao Paulo; Departamento de Molestias Infecciosas e Parasitarias da Faculdade de Medicina da Universidade de São Paulo; Faculdade de Medicina da Universidade Municipal de Sao Caetano do Sul; Instituto Nacional do Câncer-Rio de Janeiro; Instituto de Ensino e Pesquisa do Hospital Sírio-Libanês; Vital Strategies; Weill Cornell Medicine, USA

## Abstract

**Objectives:** To evaluate if antibodies induced by infection with a different SARS-CoV-2 virus strain neutralize the P.1 variant.

**Methods:** Convalescent sera from 60 individuals following a documented SARS-CoV-2 infection were assayed for neutralizing antibody titer against both strains.

**Results:** Fifty-six and 50 sera were positive for neutralizing antibodies against the ancestral and P.1 strains, respectively. Neutralization titers were higher against the ancestral strain, but in the majority of patients differences did not differ by more than a single dilution.

**Conclusions:** Neutralizing antibodies that were generated following infection with SARS-CoV-2 B.1.1.28 were effective *in vitro*, against the SARS-CoV-2 P.1. variant.

## Introduction

Since the beginning of the coronavirus disease 2019 (COVID-19) pandemic, one of the major concerns has been the duration of immune protection and its specificity following the initial infection. The long term clinical and immunological consequences of anti-viral antibody production against the infecting strain remain unclear and correlations between antibody levels and protection against re-infection by SARS-CoV-2 variants remain under-reported^1,2^. Neutralizing antibodies are antibodies that react with surface components of SARS-CoV-2, specifically the spike protein, and prevent the virus from interacting with specific receptors on target cells and thereby initiating a productive infection^1,3^. Measuring and comparing the neutralization capacity of antibodies in sera from convalescent individuals previously infected with SARS-CoV-2 strains circulating at the beginning of the pandemic with genetic variant strains present at late pandemic stages will provide much needed information regarding the occurrence of cross-immunity between different viral strains.

Beginning in November 2020, a novel SARS-CoV-2 variant, lineage P.1, was identified in Manaus, Brazil^4^. Genome sequencing demonstrated that this variant is characterized by 17 mutations, including three in the gene coding for the spike protein (K417T, E484K and N501Y). These mutations result in antigenic changes in the spike protein and could impair the efficacy of neutralizing antibodies that were generated against a previous SARS-CoV-2 strain^5^. The P.1 SARS-CoV-2 variant has spread throughout Brazil and has become prominent in perpetuating and expanding the pandemic in this country.

The aim of the present study was to evaluate if neutralizing antibody responses induced by infection with the SARS-CoV-2 virus that was dominant at the beginning of the pandemic remained effective when tested against the P.1 lineage.

## Methods

### Setting and Patients

Included patients were participants in *The Corona São Caetano Program*, a primary care initiative offering COVID-19 care to all residents of São Caetano do Sul, Brazil^6^. Sixty participants who were positive for SARS-CoV-2 infection between May 4 and May 16, 2020 were enrolled into this study. All were > 18 years old, had a confirmed SARS-CoV-2 infection by RT-PCR analysis of nasopharyngeal and throat swabs, performed at the Virology Laboratory at Instituto de Medicina Tropical de São Paulo. After explaining the study and obtaining written informed consent, peripheral blood was collected from each participant at one week intervals for 5 consecutive weeks (days 1, 7, 14, 21, 28) beginning up to 72 hours from the time of initial diagnosis.

### Virus Identification- RNA extraction, PCR amplification

RNA extraction was performed using the QIAamp viral RNA kit, according to the manufacturer instructions. For the RT-PCR assay, we used the commercial RealStar® SARS-CoV-2 RT-PCR Kit 1.0, (Hamburg, Germany), develop by *Altona Diagnostics*. DNA amplification was performed using the Roche LightCycler® 96 System.

### Virus isolation for virus neutralization test

An ancestral variant (EPI_ISL_1557222) which was classified as belonging to B.1.1.28 lineage, was cultured from nasopharyngeal swab taken from an infected patient from Sao Caetano do Sul, City, Brazil in April, 2020. The P.1 SARS-CoV-2 variant (EPI_ISL_1060902) was obtained from a nasopharyngeal specimen of a patient from Manaus City, Brazil, in December, 2020 that was previously classified as belonging to the P.1 lineage by virus genome sequencing ^4^. Virus isolation and titration were performed according to Araujo et al. ^7^ and Park et al^8^. with some modifications.

To isolate SARS-CoV-2 we used Vero cells (ATCC® CCL-81™). Cells were seeded in a 25 cm^2^ cell culture flask (polystyrene sterile, non-pyrogenic flask,12.5 cm^2^, 25mL, Biofil®, China) in a concentration of 5 ×10^5^ cells/mL in 3.0 mL Dulbecco Minimal Essential Medium (DMEM) supplemented with 5% heat-inactivated fetal bovine serum (FBS) (Vitrocell Embriolife, Campinas, Brazil) and incubated overnight at 37°C. The next day, the supernatant was discarded, and 0.5 mL of a homogenized nasopharyngeal (NP) swab specimen was added into the culture flask. After 1 h of incubation (adsorption), we supplemented the volume with 3.0 mL DMEM containing 2.5% FBS and 1% penicillin-streptomycin and inoculated the cultures in a humidified 37°C incubator in an atmosphere of 5% CO_2._ We observed for the presence of cytopathic effects (CPE) daily for 3-5 days. The supernatant was collected, and virus replication was confirmed through CPE and by RT-PCR.

### Virus titration

The viral titer was expressed in TCID_50_/mL and calculated using the Spearman & Kärber algorithm, as described by Hierholzer & Killington^9^ (See Supplementary Virus titration).

### Virus Neutralization Test (VNT)

The cytopathic effect (CPE)-based virus neutralization test (VNT) was adapted from Nurtop et al.^10^ and was previously described by Wendel et al.^11^ and Mendrone-Junior et al.^12^. VNT was performed with SARS-CoV-2 (EPI_ISL_1557222 and EPI_ISL_804814) in 96-well microtiter plates containing 5 × 10^4^ Vero cells/mL. Vero cells were seeded in a 96-well microtiter plate and allowed to grow for 24 hours prior to infection. Sera to be tested were heat-inactivated for 30 minutes at 56°C. Then, 50 μL of twofold serially diluted sera from 1:20 to 1:2560, were added mixed vol/vol with 10^3^ TCID_50_/mL of SARS-CoV-2 and incubated at 37°C for 1 h for virus neutralization. The sera - virus mixture was transferred onto the confluent Vero cell monolayer and incubated for 72 hours. Cultures at 37°C and 5% CO2 were observed daily for a CPE. After 72 hours, the plates were analyzed by light microscopy (Nikkon, Japan), distinguishing the presence/absence of CPE-VNT. To verify the initial observations after 72 hours the monolayers were fixed and stained with Naphthol Blue Black (Sigma-Aldrich Co., Deisenhofen, Germany) dissolved in sodium acetate-acid acetic for 30 minutes. Dilutions of serum associated with CPE were considered as a negative result. The absence of CPE or a complete neutralization of SARS-CoV2 inoculum was considered as a positive result. Consequently, the VNT was the highest dilution of serum that neutralized viral growth (absence of a CPE).

For each reaction we used as a positive control diluted virus in DMEM with 2.5% FBS and as a negative control only DMEM with 2.5% FBS. In addition, a serum control was a serum specimen taken from a patient with a SARS-CoV-2 infection in São Paulo with a known VTN titer. The antibody titer was calculated as the highest dilution where CPE was completely inhibited. Titers >1:20 were reported as positive. Virus isolation and VNT were performed in a Biosafety Level 3 laboratory.

### Statistical analysis

We investigated whether convalescent individuals have similar serum virus neutralization titers against isolates of SARS-CoV-2 lineage “P.1” as compared to the ancestral lineage. Neutralization titers were grouped by weeks (1=1-7, 2=8-14, 3=15-21, 4=22-28, 5=29-35, 6=36-42 days after onset of symptoms), as most patients were visited and had their sample collected only once per week. For the few occasions when patients had two visits during the same week, the maximum value of the two was considered.

The percentage of positive results (neutralization titers ≥ 20) are presented by week after the onset of symptoms, together with their 95% binomial confidence intervals. Continuous variables were summarized as median and interquartile ranges and presented in box plots.

The equality of matched pairs of observations was tested using the Wilcoxon matched-pairs signed-rank test. We also calculated the paired differences in dilution titers and subsequently categorized these differences into “≤ -2”, “in between -1 and 1” and “≥ 2”.

Database management and statistical analysis were performed using the Stata-15 software (Statacorp, College Station, Texas, USA).

## Results

Demographics and clinical characteristics of the study population are presented in Table 1 Supplementary. The majority (58.3%) were female and 46.6% were <40 years old. All had mild or moderate disease with symptoms lasting a median of 3 days; none required hospitalization.

**Table 1.** Summary of neutralization titers against each lineage, by week since onset of symptoms.

| **Weeks since onset of symptoms** | **Type** | **N** | **Median (p25-p75)** | **p-value\*** |
| --- | --- | --- | --- | --- |
| **1** | Ancestral | 22 | 0 (0-0) | 0.5275 |
|  | P.1 | 22 | 0 (0-0) |  |
| **2** | Ancestral | 50 | 80 (0-160) | 0.0008 |
|  | P.1 | 49 | 20 (0-80) |  |
| **3** | Ancestral | 55 | 160 (80-640) | 0.0010 |
|  | P.1 | 54 | 80 (20-320) |  |
| **4** | Ancestral | 54 | 160 (80-640) | 0.0011 |
|  | P.1 | 52 | 80 (20-240) |  |
| **5** | Ancestral | 52 | 320 (80-640) | 0.0017 |
|  | P.1 | 49 | 80 (20-160) |  |
| **6** | Ancestral | 35 | 320 (80-640) | 0.0157 |
|  | P.1 | 34 | 120 (40-320) |  |

Signed-rank tests were used to compare differences in the matched pairs.

Fig. 1 demonstrates the rapid increase in neutralization titer against the original viral lineage over the initial three-week period, followed by a plateau at or near the maximum level over the subsequent three weeks. The increase in neutralization titer against the P.1 variant closely paralleled what was observed with the ancestral lineage, but the levels were on average about 10% lower at every timepoint. Four patients (6%) did not have evidence of neutralizing antibodies against both the ancestral lineage and P.1.lineage, while an additional 6 patients were negative for neutralizing antibodies only against the P.1.lineage throughout the study period.

**Figure 1.**
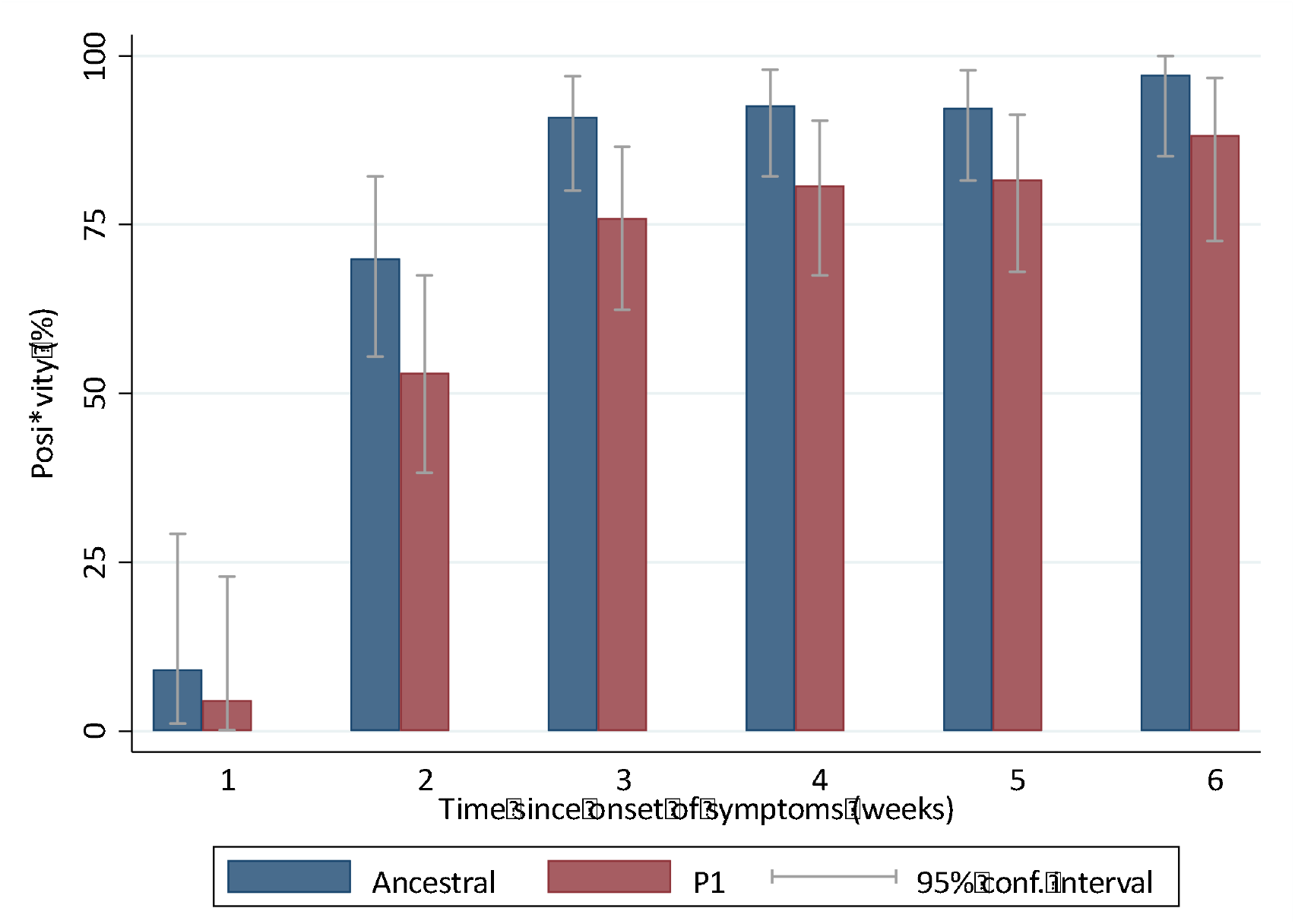
Positivity of neutralization titers against isolates “ancestral” lineage compared to those against the “P.1” lineage, by week since onset of symptoms.

The neutralization titers against the ancestral and P.1 strains at each week of testing are shown in Table 1. Median values were consistently higher against the ancestral strain than against the P1 lineage. All differences reached statistical significance, except for week one.

When comparing the neutralization titers against the ancestral strain and P.1, we observed that this difference was not more than a single dilution in 57.8%, not more than two dilutions in 37.6% and > 2 dilutions in only 4.6% of the sera (Table 2 Supplementary material). The differences remained fairly constant over the time course of the study (Fig. 2 Supplementary material).

## Discussion

The great majority of patients infected with one SARS-CoV-2 strain (B.1.1.28 lineage) developed neutralizing antibodies that were also effective *in vitro*, although at a slightly reduced titer, against the P.1 variant, without having a prior exposure to this strain. It is likely that the small difference in neutralization titers would not diminish the ability to effectively prevent or minimize infection following exposure to the P.1 variant. Thus, previous infections with the ancestral strains will apparently provide a sufficient degree of protection from the P.1. variant in the majority of previously infected individuals. This protection may also extend to other components of the immune response, such as long-lived memory T cells^13^. It has already been postulated that even when neutralizing antibody levels have dropped below a detectable threshold, immune memory could lead to induction of an anamnestic antibody response following re-exposure to the same or different strains of the virus, and this is likely to be protective against severe disease^1^.The reason(s) for the difference in neutralization antibody titers against the ancestral lineage that produced the infection and the P.1variant remain to be determined. It is likely that they reflect variations in the conformation of the spike protein that influences the efficacy of antibody binding^5,14^. This possibility has been evaluated in only a few studies^5^ and the mechanism remains unresolved.

It is important to acknowledge limitations of our study. First, our cohort included a relatively small number of patients, all of whom recovered from mild or moderate episodes of COVID-19. It would be of interest to expand the study to determine if individuals with more severe disease differed in the occurrence or titer of antibodies cross-reactive to P.1.

Secondly, our study only assessed neutralizing antibody titers for the first six weeks after initial infection. The length of time that the neutralizing antibody titers to the initiating strain and to the P.1 variant are maintained remains to be determined. Extending this study to later time points would be valuable to gain this additional information. Data on humoral immunity to other human coronaviruses have indicated that antibody levels wane over time^2^. A few studies have assessed antibody titers to MERS CoV and SARS CoV in the months and years following primary infection. Although limited in size these studies indicated that total binding antibodies and neutralizing antibodies progressively decreased such that by 2–3 years all infected individuals had minimal detectable antibody responses. There are reports of reinfection with homologous coronaviruses after as little as 80 days^2^.

Thirdly, we did not measure other components of the immune response, such as cell-mediated immunity, which would further contribute to viral neutralization *in vivo*. It is likely that large prospective epidemiologic studies will be required to establish the role of neutralizing antibodies in preventing infection, or minimizing their consequences, by SARS-CoV-2 variants in previously exposed individuals. This will also most likely vary depending on the extent of changes in spike protein antigenicity between the infecting strain and the specific variant being analyzed.

In conclusion, our data provides the encouraging information that a previous SARS-CoV-2 infection likely provides a substantial degree of protection against the P.1 variant, even in most individuals with a relatively mild infection.

## Data Availability

Raw data were generated at Instituto de Medicina Tropical de Sao Paulo, Brazil. The data that support the findings of this study are available on request from the corresponding author.

http://www.imt.usp.br/

## Ethics

The study was approved by the local ethics committee (CONEP, protocol No. **CAAE:** 30419320.7.0000.0068, dated April 18, 2020) and all subjects provided informed written consent.

## Acknowledgment

We thank the staff from Laboratorio de Investigacao Medica em Virologia (LIM52)-Instituto de Medicina Tropical de Sao Paulo for general laboratory support.

We also thank Prof.Ester Sabino from Instituto de Medicina Tropical de São Paulo, for providing the isolates from nasopharyngeal specimens from Manaus City, Brazil.

## Author contributions

All authors made a significant contribution at different stages of the study and contributed to data interpretation. MCMC and SSW wrote the first draft of the paper. ALB was responsible for the statistical analysis. All authors participated in writing subsequent drafts. All authors approved the final version of the manuscript.

---

**Table. 1 Supplementary material:** Demographics and clinical characteristics of the studied population

| **Characteristics** | **Overall N=60** |
| --- | --- |
| Female sex, n (%) | 35 (58.3%) |
| Age, years (range) | 18-75 |
| < 40, n (%) | 28 (46.6%) |
| >40 <61 n ( %) | 23 (34.4%) |
| ≥ 61, n (%) | 9 (15%) |
| Hospitalization, n (%) | 0 |
| Duration of symptoms (days), median (range) | 3 (1-10) |

**Table 2 Supplementary material.** Paired dilution differences of neutralization titers against isolates of SARS-CoV-2 “ancestral” lineage compared to those against the “P.1” lineage, by weeks since onset of symptoms.

| Differences in dilutions | Time since onset of symptoms (weeks) |  |  |  |  |  | Total |
| --- | --- | --- | --- | --- | --- | --- | --- |
|  | N (%) |  |  |  |  |  |  |
|  | 1 | 2 | 3 | 4 | 5 | 6 |  |
| ≤ -2 | 2<br>(9.1) | 17<br>(34.7) | 24<br>(45.3) | 22<br>(43.1) | 21<br>(43.7) | 10<br>(30.3) | 97<br>(37.6) |
| -1 / 0 / 1 | 20<br>(90.9) | 29<br>(59.2) | 26<br>(49.1) | 27<br>(53.0) | 24<br>(50.0) | 22<br>(66.7) | 149<br>(57.8) |
| ≥ 2 | 0<br>(0.0) | 3<br>(6.1) | 3<br>(5.6) | 2<br>(3.9) | 3<br>(6.3) | 1<br>(3.0) | 12<br>(4.6) |
| Total | 22<br>(100.0) | 49<br>(100.0) | 53<br>(100.0) | 51<br>(100.0) | 48<br>(100.0) | 33<br>(100.0) | 258<br>(100.0) |

**Figure 2 Supplementary material.**
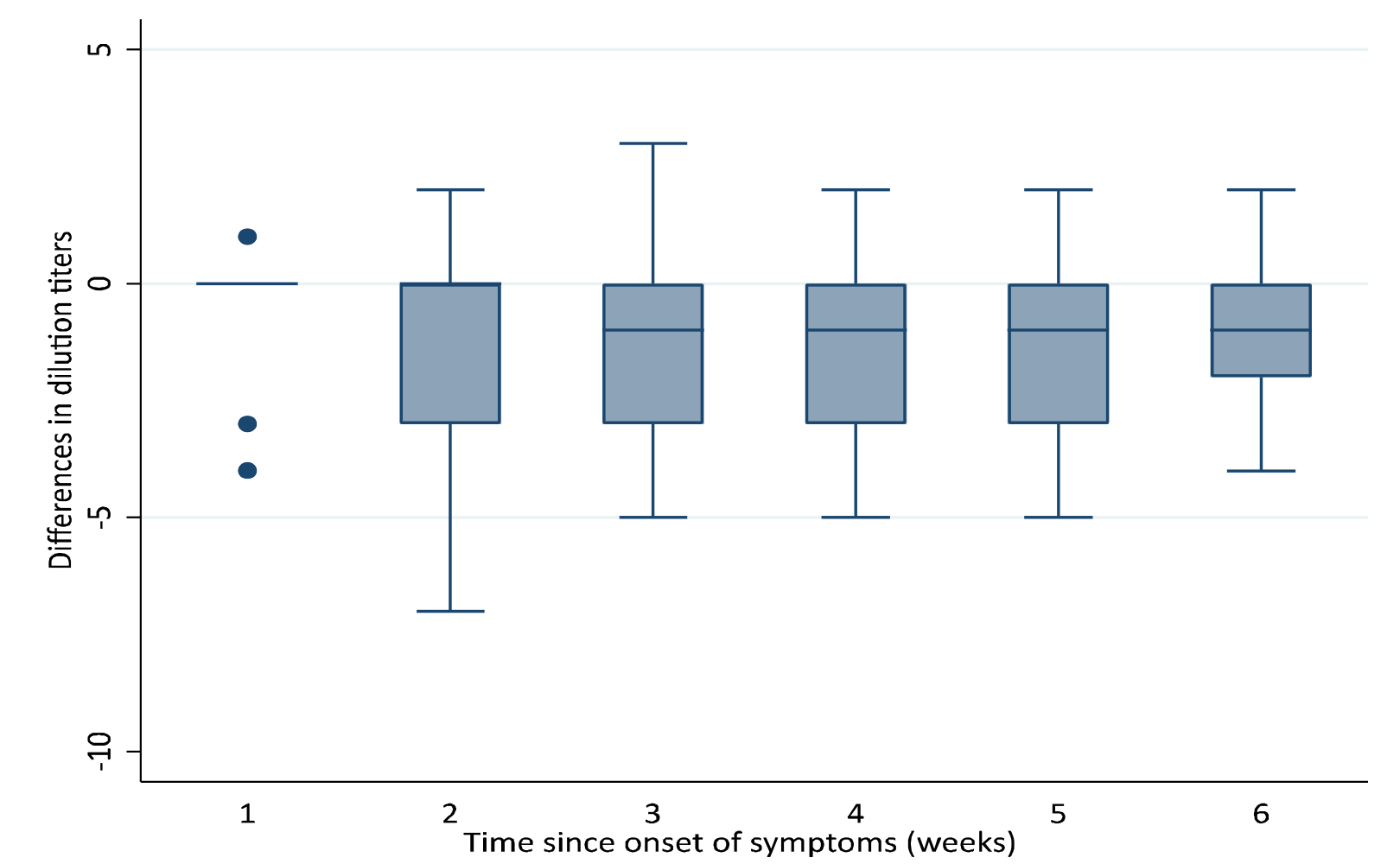
Differences in neutralization titers against the SARS-CoV-2 ancestral strain and the “P.1” lineage over the six-week study period.

## *Virus titration –* Supplementary

Twenty-four hours before viral addition, we prepared 96 well sterile polystyrene microtiter plates (Biofil®) containing 5 × 10^4^ cells/mL Vero CCL-81 cells in DMEM. A virus preparation was 10-fold serially diluted in medium (10^−1^ to 10^−12^), the original culture medium was then removed and replaced with serial dilutions of the virus in sextuplicate wells and incubated at 37°C. Observations were performed daily using an inverted light microscope (Nikon 45178, Japan) to verify the presence of CPE over a 72 h period. The monolayers were then fixed and stained with Naphthol Blue Black (Sigma-Aldrich Co., Deisenhofen, Germany) dissolved in sodium acetate-acid acetic. The viral titer was expressed in TCID_50_/mL and calculated using the Spearman & Kärber algorithm, as described by Hierholzer & Killington^1^.

